# Empiric model for short-time prediction of COVID-19 spreading

**DOI:** 10.1101/2020.05.13.20101329

**Authors:** Martí Català, Sergio Alonso, Enrique Alvarez-Lacalle, Daniel López, Pere-Joan Cardona, Clara Prats

## Abstract

Covid-19 appearance and fast spreading took by surprise the international community. Collaboration between researchers, public health workers and politicians has been established to deal with the epidemic. One important contribution from researchers in epidemiology is the analysis of trends so that both current state and short-term future trends can be carefully evaluated. Gompertz model has shown to correctly describe the dynamics of cumulative confirmed cases, since it is characterized by a decrease in growth rate that is able to show the effect of control measures. Thus, it provides a way to systematically quantify the Covid-19 spreading velocity. Moreover, it allows to carry out short-term predictions and long-term estimations that may facilitate policy decisions and the revision of in-place confinement measures and the development of new protocols. This model has been employed to fit the cumulative cases of Covid-19 from several Chinese provinces and from other countries with a successful containment of the disease. Results show that there are systematic differences in spreading velocity between countries. In countries that are in the initial stages of the Covid-19 outbreak, model predictions provide a reliable picture of its short-term evolution and may permit to unveil some characteristics of the long-term evolution. These predictions can also be generalized to short-term hospital and Intensive Care Units (ICU) requirements, which together with the equivalent predictions on mortality provide key information for health officials.

**Author summary:** Covid-19 has brought international scientific community into the eye of a storm. Collaboration between researchers, public health workers and politicians is essential to deal with this challenge. One of the pieces of the puzzle is the analysis of May 7, epidemiological trends so that both current and immediate future situation can be carefully evaluated. For this reason we have daily employed a generic growing function to describe the cumulative cases of Covid-19 in several countries and regions around the world and particularly for European countries during the Covid-19 outbreak in Europe. Our model is completely empiric and it is not using any assumption to make the predictions, only the daily update of new cases. In this manuscript, we detail the methods employed and the degree of confidence we have obtained during this process. This can be used for other researchers collaborating and advising health institutions around the world for the Covid-19 outbreak or any other epidemic that follows the same pattern.

## Introduction

An outbreak is always a challenge for the public health control systems. When the outbreak is caused by a new agent able to cause a pandemy, the challenge is even greater and should involve the whole research community as well. Globalization plays a double role in this context: on the one hand, it increases the risk for the outbreak to evolve towards a pandemic; on the other hand, sharing of data and strategies enhance the chance to control it. The new SARS-CoV-2 virus (severe acute respiratory syndrome coronavirus 2) has put the international community at the edge of a global disaster. National and local governments are working with public health agencies hand with hand in order to slow down and, eventually, control the spread of Covid-19 [1].

Daily availability of data about Covid-19 confirmed cases in different regions is a unique opportunity for basic scientists to contribute on its control by carefully analyzing dynamics and tends. In particular, mathematical models are widespread and consolidated tools to extract information of data and help making predictions [2]. Classic SIR and SEIR models (i.e., compartment models that divide a population into Susceptible, Exposed, Infectious and Recovered) are being currently employed to evaluate and predict the spreading of the epidemic episodes [3]. They have been employed in the description for example of the Ebola epidemic on [4] and [5] and in the more recent SARS epidemic on [6] among others. After the SARS epidemic on 2003, in order to account the control efforts of the governments, some modifications were introduced on the SEIR model to evaluate control measures [7,8]. Furthermore, the analysis of SEIR models have been already employed for the modeling of the Covid-19 spreading in China in a effort to fit the characteristic values [9,10]. However, during the development of the epidemic, strong control measures are luckily conditioning such dynamics and thus hindering the soundness of this approach. Furthermore, SIR and SEIR models are regulated by the limitation of the susceptible population, constrain, which in the case of Covid-19, seems to be unnecessary [11].

There is, however, another approach based on the phenomenological comparison of the curve of cumulative cases with a typical function for growing processes. Evaluating the trend of the curve during the last days, allows the future short-time behavior tendency [12]. In fact, the use of a growing function has some important advantages. Typically, the first growing function chosen is the Verhulst equation [13] which is the solution of the logistic population model and the generalization of the model [14,15] or the Richards model [16] which have been employed in several epidemic spreading like smallpox, influenza, Ebola, among others [14]. Some of these types of dynamical phenomenological growth models to study epidemic outbreaks have been compared in the initial phases of the Covid-19 epidemic for the short-term forecasting [17].

A similar growth model is the Gompertz function [18] where the main difference is the substitution of the saturation of the growing factor, linear for the Verhulst equation and non-linear for the Richards and generalized Verhulst model, by an exponential decrease. These functions are similar and they have been used in the description of epidemics and in particular have been evaluated for studying different epidemic episodes [19,20]. While the Logistic equation produces a symmetric bell-shaped function for the new cases, the Gompertz model gives rise to an asymmetric function with a fast growth of new cases combined with a slow decrease which is more similar to the distribution of new cases observed in different countries during the spreading of some epidemic. We show in this manuscript that the asymmetric nature of the Gompertz model is the proper framework to study epidemics where control measures are at the heart of its evolution since it captures the dynamic nature of the variation due to social distance measures.

Here, we employ the Gompertz growing function to analyze the dynamics of the spreading of Covid-19 in several countries to make short-time predictions of the new cases for the successive days. We forecast the dynamics of the pandemic in a similar fashion of the forecasting done previously with the Verhulst equation and the Richards model for Ebola epidemics [21]. The methodology and the results here discussed were employed for the writting of daily reports [22] and the prediction of cases for hospitals and intensive care units (ICUs).

The manuscript is organized as follows, first the methods and particularly the Gompertz function is described and the main approaches are discussed, second, the main results using this function to study the evolution of the Covid-19 in different countries are shown, and finally the conclusions of this research are detailed.

## Materials and methods

First we describe the function employed for the fitting of the data and next describe the evaluation of the errors associated to these calculations.

### Short review on Gompertz equation

We employ the Gompertz model for growing processes for the modeling of the cumulative cases of Covid-19. Such equation was originally proposed as a means to explain human mortality curves [18], and it has been further employed in the description of growth processes, for example, growing of bacterial colonies [23] and tumors [24]. The Gompertz equation reads:

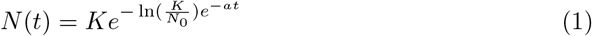

where the parameter *K* corresponds to the final number of cases, *N*_0_ is the initial number of cases for the definition of the origin of time, and parameter a is the rate of decrease in the initially exponential growth, see curves in Fig. 1(A) for different values of *a*. For the initial stage, corresponding to small times, the eq.(1) reduces to an initial exponential growth *N* = *N*_0_*e*^*µ0t*^ with rate

**Figure 1.**
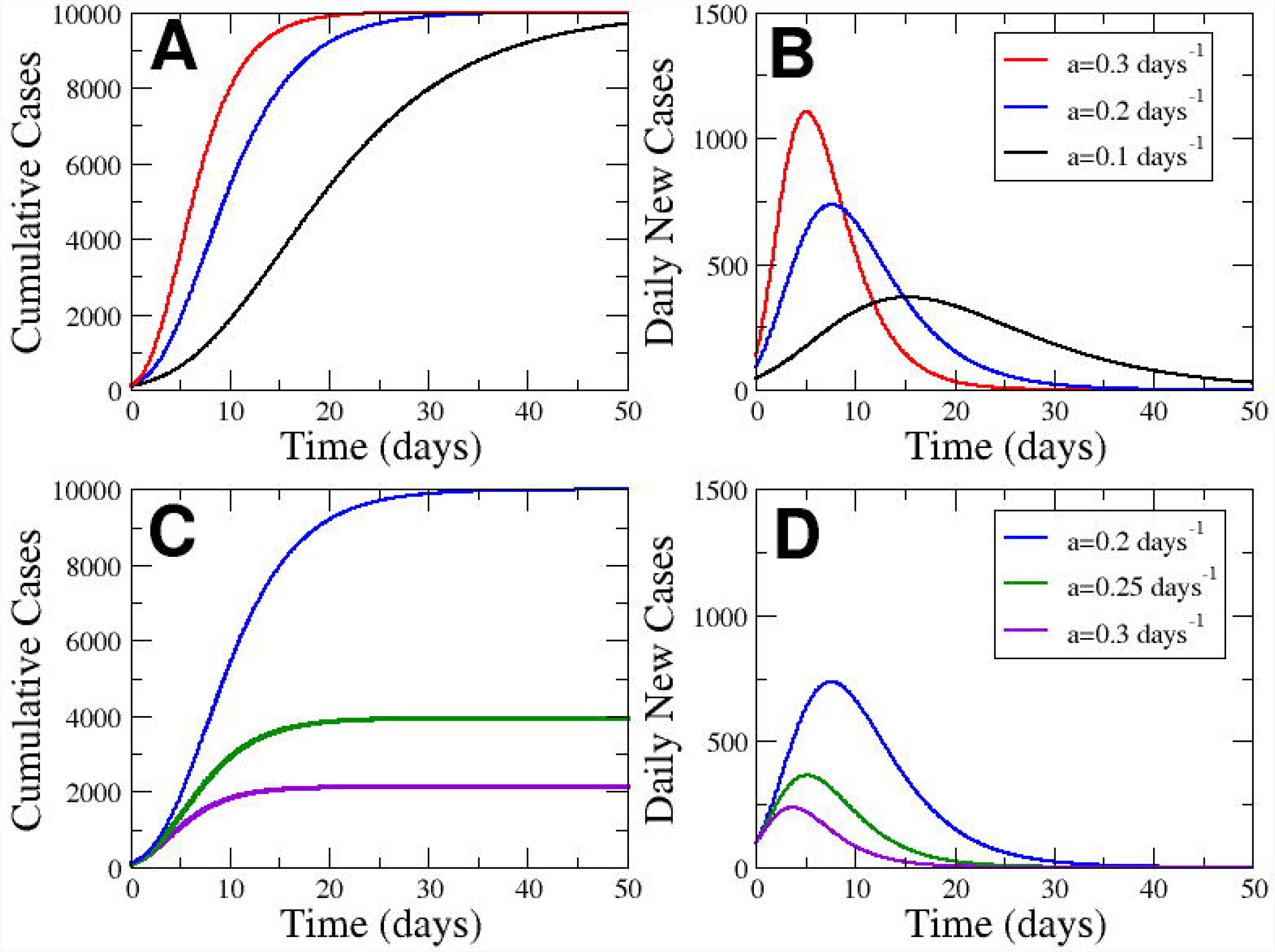
**Properties of Gompertz function**. Evolution of the cumutated cases (A) and the new cases (B) in front of time keeping *K* = 10^4^for three different values of a. Evolution of the cumutated cases (C) and the new cases (D) in front of time keeping *µ*_0_ = 0.92 for three different values of *a*.

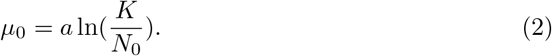

After a characteristic time the growing begins to curve till the asymptotic final value given by the saturation parameter *K*. To compare with the cumulative cases of Covid-19 we begin to measure above cases, which determines the value of *N*_0_ = 100. The exponential rate µ_0_ provides us with the relation between the parameters *K* and *a*. Therefore, we can also show the Gompertz model as function of the parameter *µ*_0_ as:

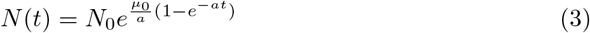

which is completely equivalent to eq.(1), however without the introduction of the total final value of total cases *K*.

On the other hand, the Gompetz function can be interpreted as the solution of the next couple of ordinary differential equations:

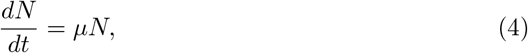

which corresponds to an exponential growth with a growing rate which is not constant but depends on the time, it decreases exponentially with time:

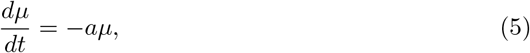

with a decrease rate a. The initial value of *µ*(*t* = 0) = µ_0_ determines the initial exponential growing of N. However, the continuous decrease of the growing rate *µ* permits the lost of the exponential growth till a complete saturation when *µ* is close to zero.

The Gompertz function shows the cumulative cases. Therefore the temporal derivative of the cumulative cases is basically the new cases, if we perform the temporal derivative:

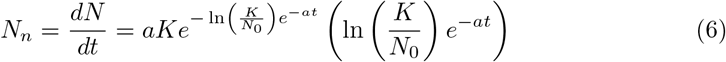

which dynamics as function of time is plotted in Fig 1(B),

Fixing the total values of cases (*K* = 10^4^) we can study the effect of a rapid decay of the growing rate, related with a large value of *a* with a more soft decrease, determined by a low value of a. See Fig. 1(A) for a visual inspection of the effect of this parameter a. The increase in the parameter *a* produces a delay of the growing process and the delay of the peak, see Fig. 1(B), where the area of the curve is constant because of the conservation of the final value K. However, in Fig. 1(C) we fix the initial exponential growth determined by µ_0_ and increase the parameter a, which decreases the final value of total cases. The amplitude of the peak is decreased by the increase in the rate *a* when the initial growth is fixed, see Fig. 1(D).

We see a maximum of the new cases in Fig. 1 which position can be calculated because it corresponds to the change from second derivative from positive to negative and it implies that

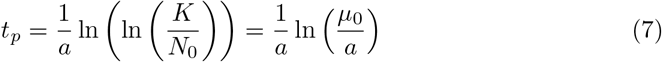

which clearly marks the effect of the parameter a. The biggest the value of *a*, the faster the appearance of the peak of new cases see Fig. 1 (B and D).

### Evaluation and propagation of errors

The fitting of the Gompertz function to the data is done with a matlab routine using the minimum least squares method. It allows the evaluation of the constants of the theoretical equations which better fits the data. Furthermore, the method also provides the error associated to these values of the constants.

The propagation of the uncertainty or error from the parameters fitted from the data to the calculation of other predictions based on the explicit values of the fitted parameters can be done using the classical methods of propagation of errors [25]. In short, if we have a quantity *U* which depends on two magnitudes *U* = *U*(*a, b*) and these magnitudes have their uncertainties a ± δa and b ± δb, if we assume the quantities are uncorrelated we can calculate the uncertainty of the new quantity as:

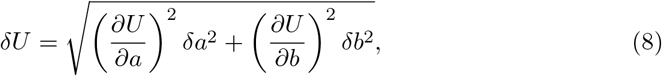

expression which is employed for example for the calculation of the time to peak, see eq.(7) and for the calculation of the 90% of the expected value of *K*. For example, for the calculation of *t_p_*, we would calculate the dependence on the parameters *a* and *K*:

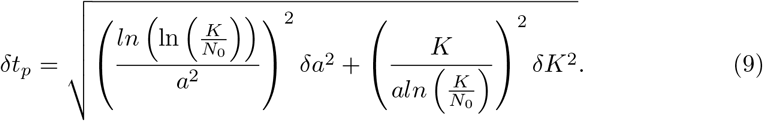

## Results

We can perform some predictions using the Gompertz function to fit the cumulative cases of Covid-19 in different countries where the epidemic is enough developed. Next we show such predictions and the main applications of the Gompertz model for the characterization of the epidemic.

### Gompertz model fits the number of cases for recovered regions

Gompertz model [26] correctly describes the trend of the cumulative confirmed cases in most of regions when fitted to epidemiological data, see Fig. 2. Despite the empirical essence of Gompertz model, it is able to quantify the observed dynamics and to predict short-term evolution. This way of quantifying observed trends is an interesting tool to objectify observations of the cumulative cases of Covid-19 and to contribute on the evaluation of the control interventions that are being implemented in each region, note for example the effect of parameter a in Fig. 1(D).

**Figure 2.**
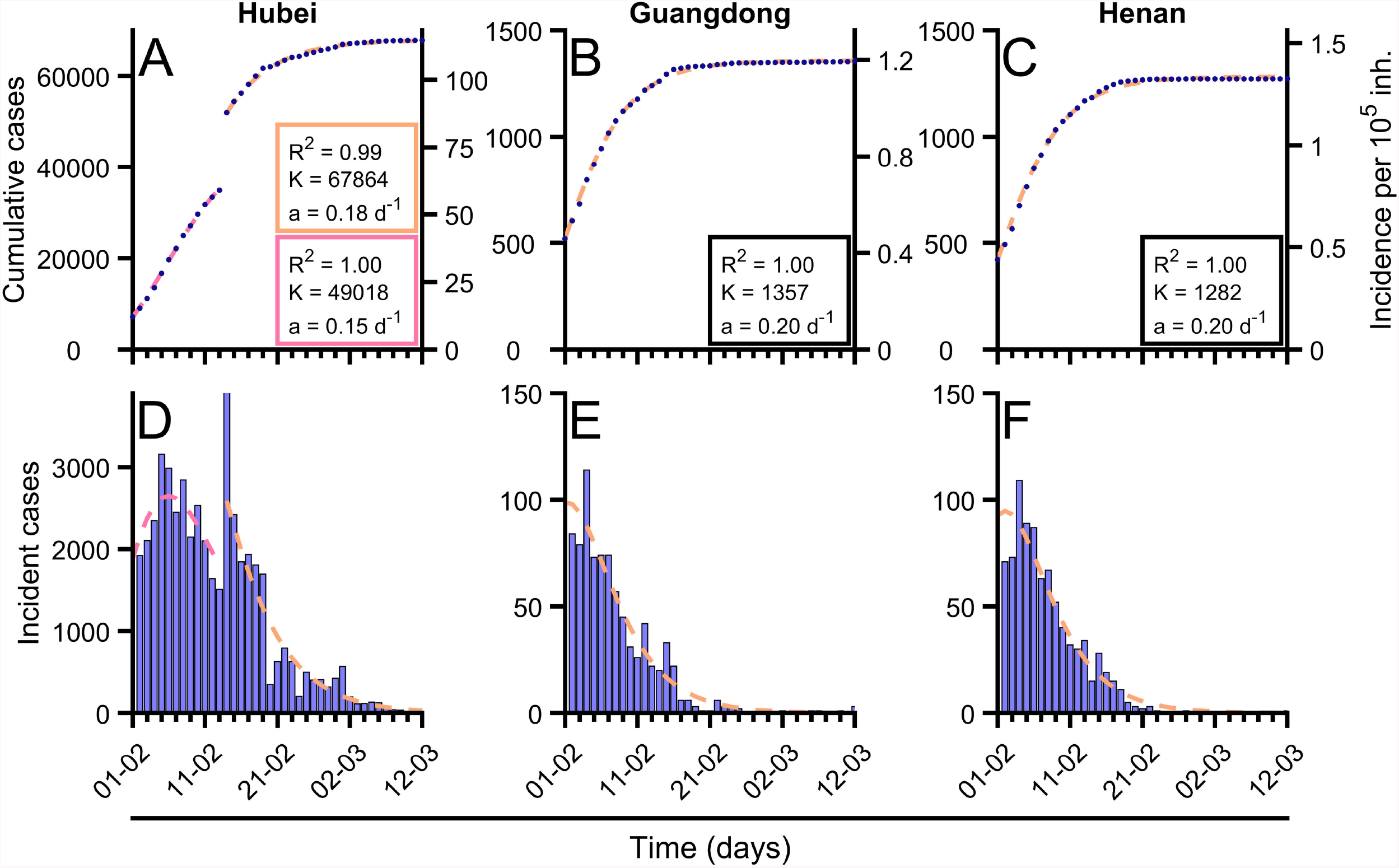
**Fitting of Gompertz function to the cumulative confirmed cases of Covid-19 in different countries.** (A-C) Evolution of total confirmed cases in different regions of China (blue dots) and fitted Gompertz function in each region (orange dashed line). In Hubei case (A) as there is a sudden change in reporting criterion there were fitted two Gompertz adjustments, previous change (pink dashed line) and post change (orange dashed line). (D-F) Evolution of incident new cases in different regions of China (blue bars) and fitted Gompertz function in each region (orange dashed line). In Hubei case (A) as there is a sudden change in reporting criterion there were fitted two Gompertz adjustments, previous change (pink dashed line) and post change (orange dashed line). The obtained values of parameter a (related with growth rate), *K* (final number of cases) and mean-squared error (R^2^) are shown for each of the fittings. Data were updated on March 5th from [27].

We perform a systematic analysis of the dynamics of the cumulative cases of Covid-19 in different regions where the spreading of the epidemic finished in China, see for example the three examples in Fig. 2 where the Gompertz function has been fitted. The Gompertz function, see eq.(1), successfully reproduces the growing of the cumulative cases for the different regions in China. Note, however, that the the fit in Hubei is divided in two regions because a change on the protocol of reporting cases. The new cases are also successfully fitted, see three panels below in Fig. 2, with the corresponding function derived of the Gompertz model, see Eq.(6). To fit the function to the data we have evaluated the values of the fitting parameters *a* and *K*, which accompany the corresponding panels in Fig. 2.

The same function with different values of the parameters *a* and *K* is employed in other countries where the epidemic is already under control. Almost all the empirical series are successfully fitted with the Gompertz function. However, there are some countries where new outbreaks appear and, therefore, the control measures are not kept constant (sudden change of parameter a) where the Gompertz function fails to conveniently fit the data. In general, although its simplicity, the Gompertz function is a good approximation for the number of cumulative cases of Covid-19 during the epidemic spreading. Given the excellent fit, this model can be applied to the final set of data to analyze and classify the epidemic characteristics of the Covid-19 given two key parameters for each one. Parameter a, which gives a measure of the type of control applied, allows for comparison among different countries which can be now assessed in a very robust manner. We will see in the next subsection that it also opens to the possibility of using this function as a quantitative empirical model to use prediction.

Let us focus now on this classification according to control measures. We show in Fig. 3 the values resulting for the fitting of the Gompertz function to the data from several regions in China. Assuming that the measures of control done in China were considered very restrictive, we can assume that the values obtained in these regions and shown in Fig. 3(A) are the upper limit of the parameter for other countries. The actual value obtained is around a = 0.2 days^−1^.

**Figure 3.**
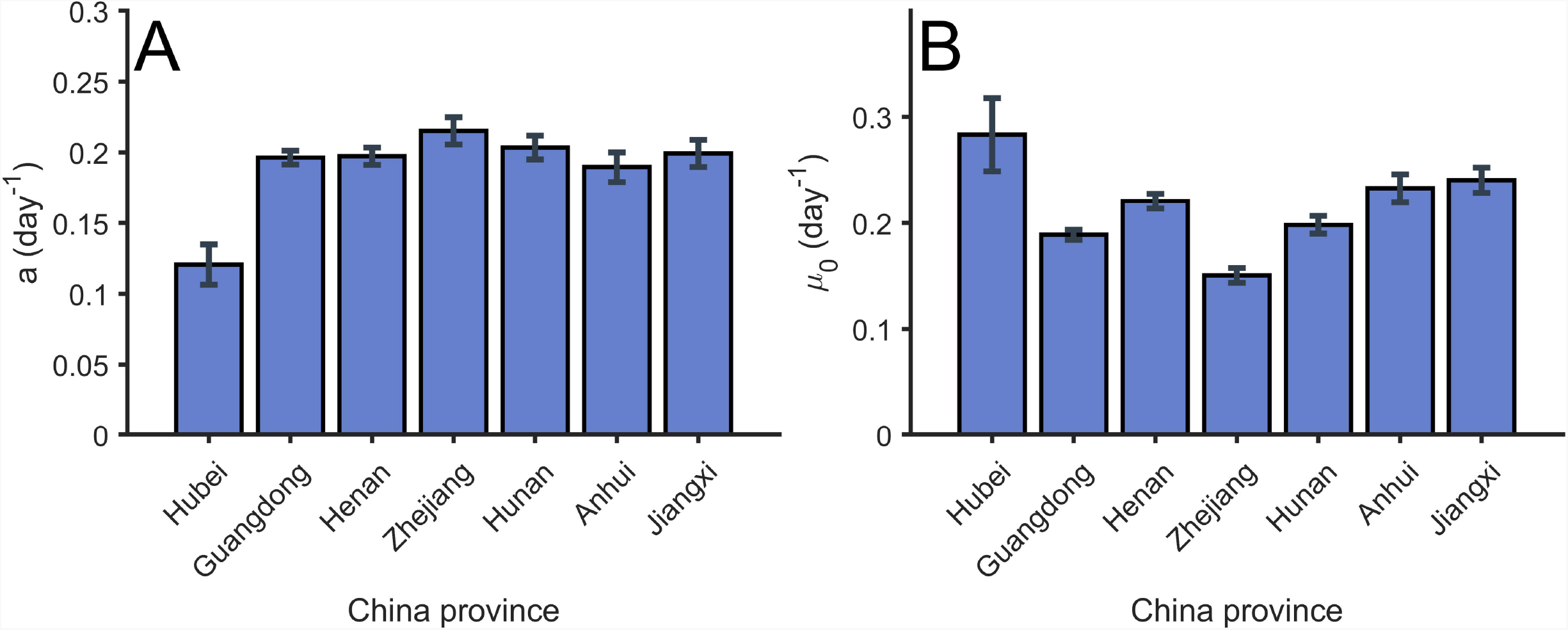
**Values of parameter *a* and *µ*_0_ of the Gompertz function in different regions in China.** (A) Value of the parameter *a* obtained from the fitting of the total confirmed cases in several regions in China. Error bars parameters confidence intervals of level α = 0.01. (B) Value of the parameter *µ*_0_ obtained from the fitting of the total confirmed cases in several regions in China. Error bars parameters with confidence intervals of level α = 0.01.

Furthermore, we can evaluate the value of the parameter µ_0_ for the initial exponential growth of the different regions, see details on Fig. 3(B). We obtain similar quantities in all the regions in China and it informs about the growing rate of the epidemic in China, which characteristic time scale is in this case similar to the decreasing rate a calculated above.

### Short-term predictions are obtained from Gompertz model

Although the understanding of the epidemic from the final picture of the dynamics is a valuable result for the treatment of future epidemics, the main goal of the modeling of epidemics is the actual possibility of prediction of the behavior during the incidence of the epidemic. We use the Gompertz model during the epidemic episode of Covid-19 in several countries in Europe.

First, we evaluate the predictions with the data obtained in the different regions in China to estimate the error of the fitting procedure of the Gompertz function before the saturation of the number of cases. We begin with the first day after 100 cumulative cases of Covid-19 and we successively fit a Gompertz function to the previous values of cumulate cases to estimate the values of parameters a and µ which will permit the estimation of the values for the cases for the next days. In Fig. 4(A-C), we show the fitting of the Gompertz function to the values of cumulative cases at three different times. The fittings of the function at different times differ with the final values of the total function shown in Fig. 2(D,E) and therefore, the values of the three fittings produces different values of the parameters a and µ_0_. However, the evolution of the values converges to the global fitting of the function to the whole set of data, see Fig. 3(D-F).

**Figure 4.**
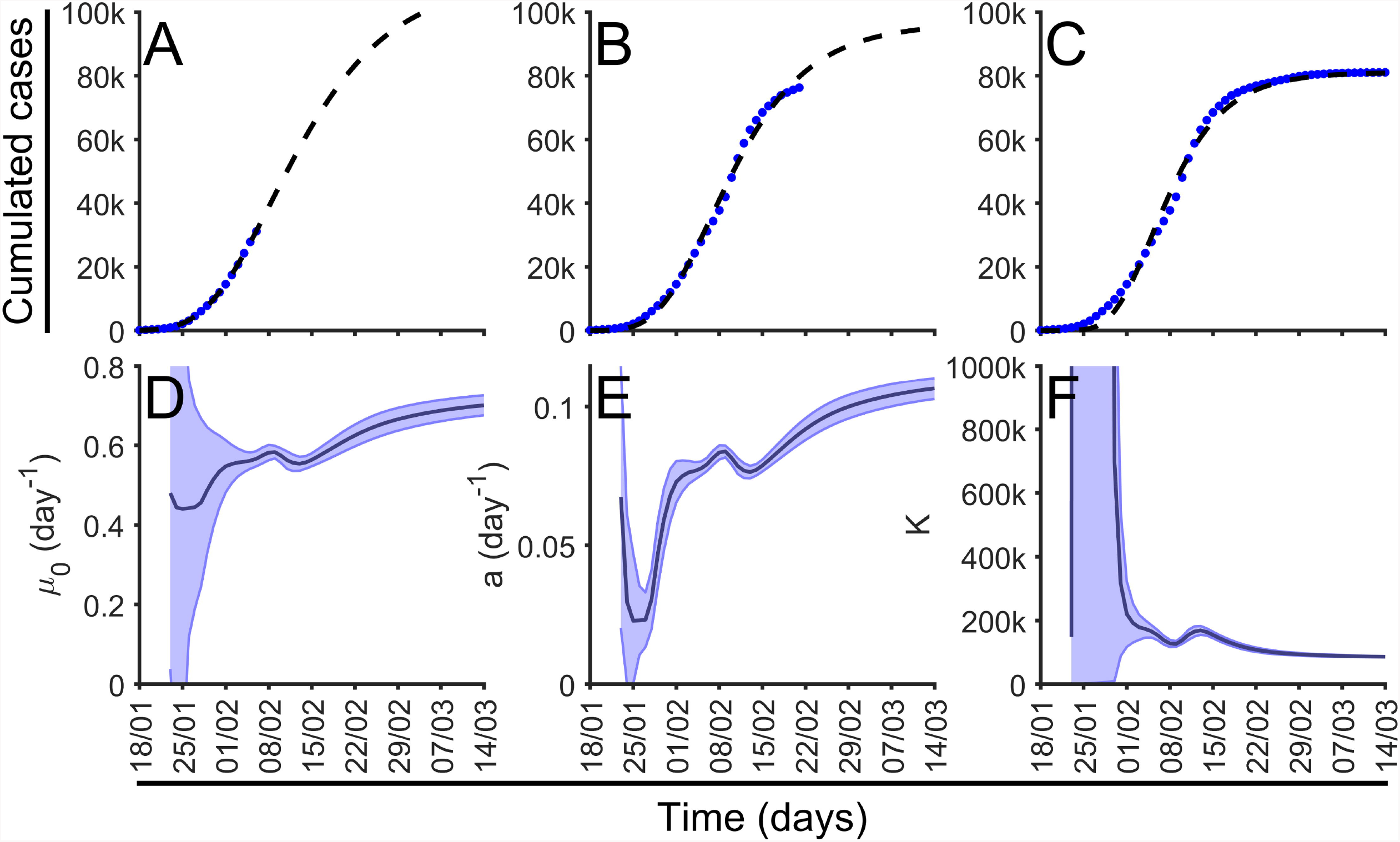
**Dynamical fitting of Gompertz function and parameters evolution.** (A-C) Gompertz fitting for China at three differents time points, february, February and March. Number of cumulative cases is shown in blue dots together with the function fitted (black dash line). (D-F) Dynamic calculation of parameters, *a* and *K* in dark blue, light blue mark error bars parameters with confidence intervals of level α = 0.01.

It is clear that the values of µ_0_ and parameters *a* and *K*, initially vary till they stabilize to a certain value at the end of the set of data. Such large variations show clearly the long-time predictions are complicated. However, we can perform short-time predictions for the number of new cases if we extrapolate to the near future the Gompertz function with the updated values of a and µ_0_ for the cumulative cases. We have systematically extrapolated the new cases for each temporal data of the series of cumulative cases of Covid-19 in the different regions in China and we have obtained a successful agreement of the predictions with the actual data for the whole series, see below for more extensive results taken into account a large number of countries.

### Short-term predictions can be applied to ongoing epidemics

The epidemic is still spreading along Europe and we have been fitting the Gompertz function to the total cumulative cases during the last two months. Most of the countries have already arrived to the saturation stage and the fitting of the function allows the evaluation of the control measures. See the examples shown in Fig. 5, where a Gompertz function satisfactorily fits the existing data. Note that Gompertz function is able to fit countries at different epidemiological phases. We have systematically assessed short-time predictions for all European countries, United Kingdom, Norway and Switzerland everyday since March 17th [28] as well as for Spanish and Italian regions [29].

**Figure 5.**
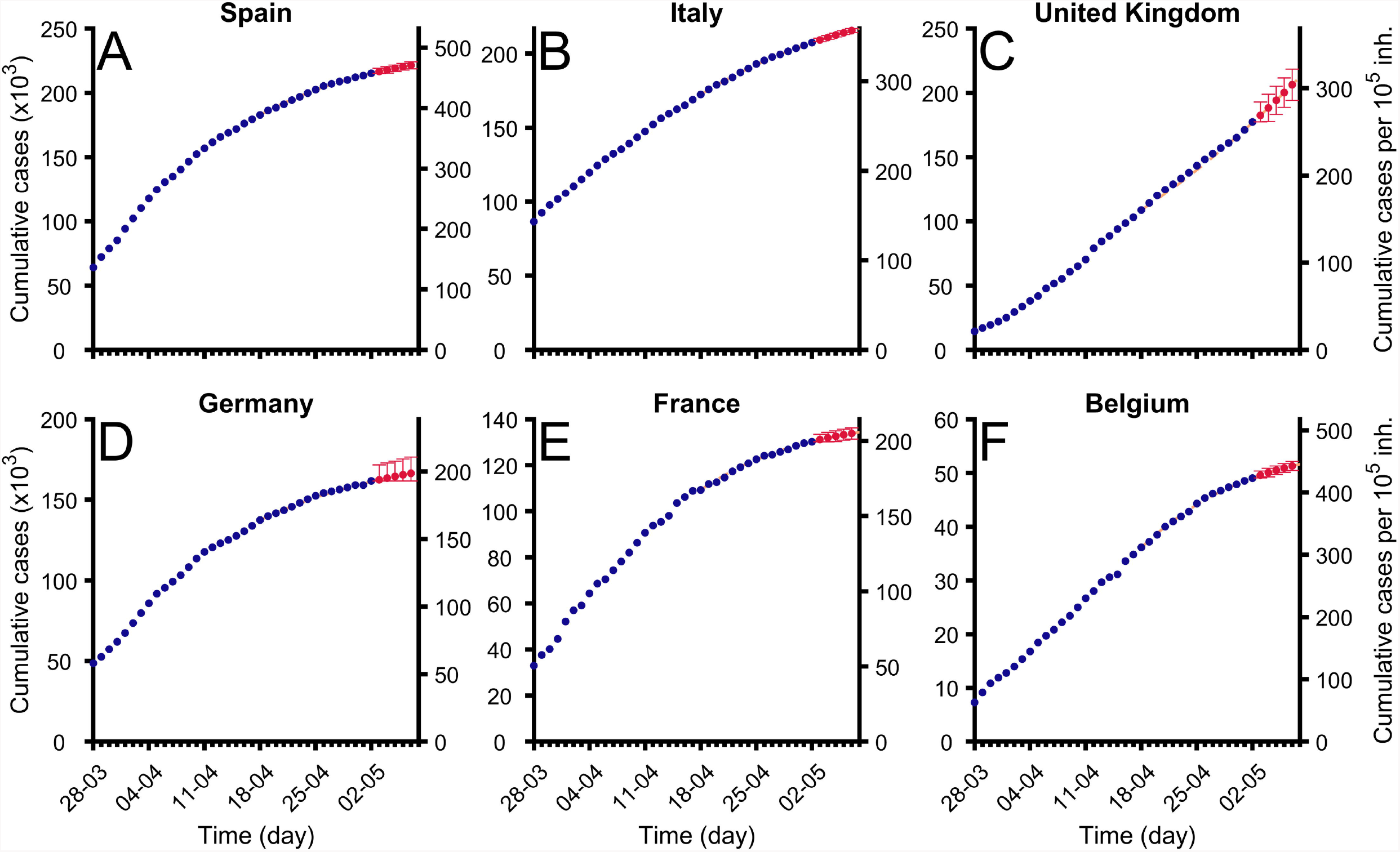
**Fitting of Gompertz function to cumulative cases in some countries in Europa.** Evolution of total confirmed cases in different regions (blue dots) and fitted Gompertz function in each region (orange dashed line). Red points show predictions for next days and error bars marks their confidence intervals of level α = 0.01. Data were updated on April 9th from [27]. (A) Spain (B) Italy (C) Germany (D) France (E) United Kingdom (F) Belgium.

Typically, the evolution of confirmed cases shows a biphasic behavior: an initial lag phase where no significant increase in the incidence is observed, which would correspond to the period where most of the cases are imported, followed by a subsequent phase where growth is evident, which would be the reflection of triggering local transmission. Gompertz model is fitted to the later phase, i.e., it is applied from the moment where a clear increase in confirmed cases is observed, typically above 100 cases to avoid the initial phase dominated by the importation of cases from other zones.

We fit the function over time to be able to predict the evolution of the cumulative cases to generate some useful information which may help the political institutions to adopt the appropriate control measures, see supplementary S1 Fig for the fitting to a selection of countries in Europe. Such fittings are based on the calculation of the values of a, see supplementary S2 Fig, and *K*, see supplementary S3 Fig, in the selection of countries.

### Evaluation of the errors in the short term predictions obtained with the Gompertz model

To evaluate the quality of the predictions we have run systematically the prediction routines along the past, for all the days of the spreading of the Covid-19 in all the countries with more than cases at April 11. The data were obtained from the ECDC [30]. The prediction evaluates the real number of cases giving rise to two different indexes: First, the average relative error of the prediction with the real quantity, and, second, if the real case is inside the error of the prediction: these two indexes allow us to calibrate the error bars of the model since we can calculate the percentage of success.

To construct the predictions we can use all the data available from the day where cumulative cases cross the threshold value of cases. However, the successive changes on the control measures may affect the parametrization of the curves. We have improved the predictions employing only the last values of the data, after the start of local transmission in the epidemics.

In Fig. 6 we show the relative error of the predictions with respect to real data. We obtain relative errors for the prediction for the next day of around 2%. The error increases for the predictions for the next days till the average error of around the 5% for the fifth day, see Fig. 6(A).

**Figure 6.**
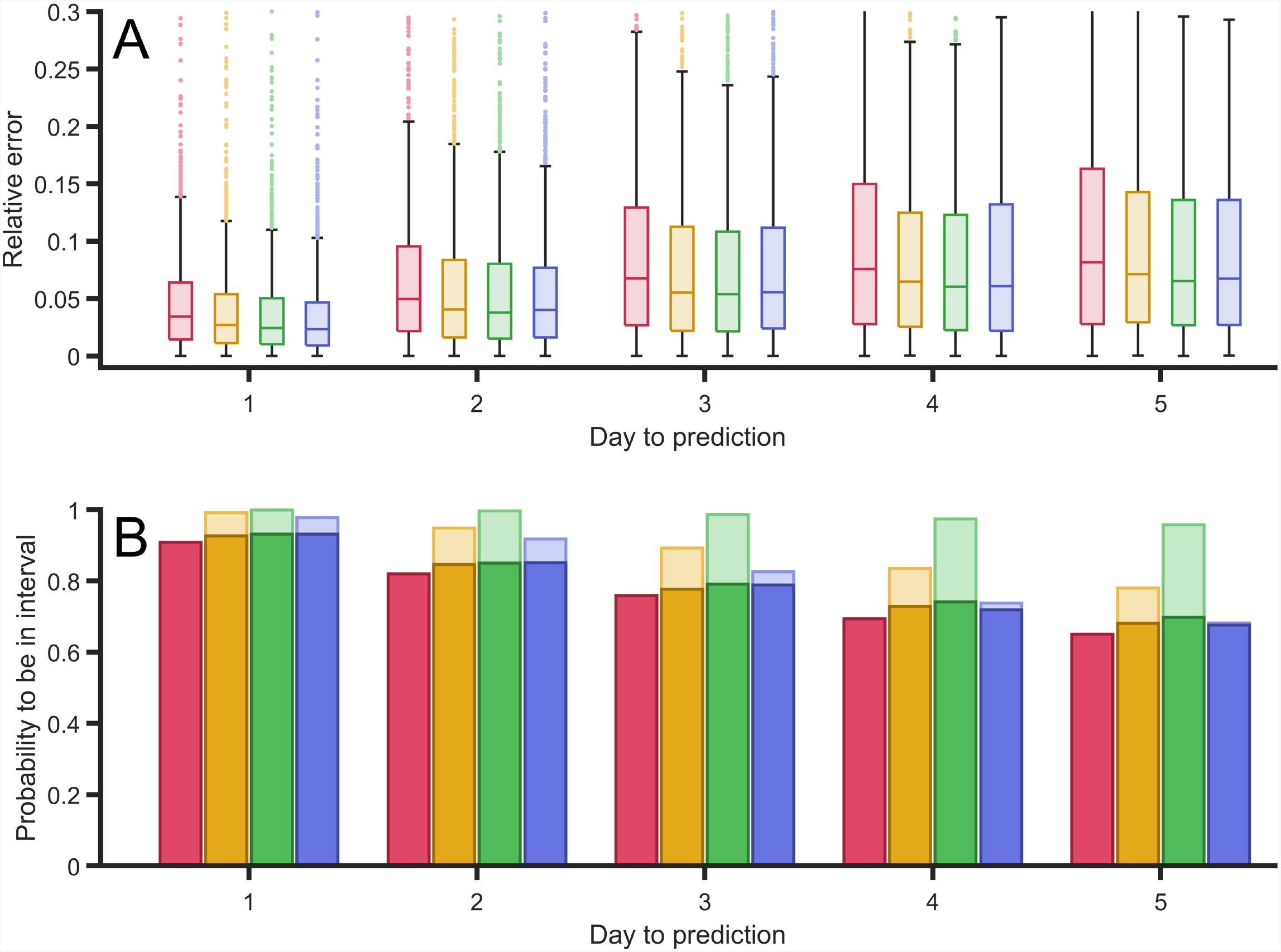
**Error of the predictions done by the dynamical fitting of Gompertz function.** (A) Relative error between the predictions of the confirmed case for the next five days, in comparison with the actual confirmed cases in several countries. (B) Probability to obtain the actual real value inside the interval of confidance inside the error bars for the next five days. These errors were computed with retrospective using all countries that had more than cases in April 9th using ECDC reported cases [30].

The predictions are obtained with a certain uncertainty due to the uncertainty on the estimation of the parameters of the Gompertz function. Therefore we evaluate in Fig. 6(B) the probability of the real value to be inside the uncertainty around the predicted value. The probability for the first day is around 90% of confidence while this probability decreases for the next days to around 60% for the fifth day, see Fig. 6(B). We certain have a great success in the predictions at short-times of the cumulative cases and therefore the new cases, and, as it was expected, the accuracy of the predictions decays with the prediction time.

### Short-term prediction error is corrected with filters

In the predictions done in the previous section at a given day, we have used the reported data from days before in order to fit the parameters of the Gompertz function. In this fit, we have given the same weight to all the days. We have given all data points the same standing. From the methodological point of view we can improve our predictions using filters to give more relevance to the last data points. This is, we can take into account more the last days before the prediction in order to improve it. This is specially useful to rapidly capture changes in trends, as for instance those that we found around the peak of new cases.

We have tried several options and concluded that three different filters must be analyzed. We proceed to show how they behave using the data sets for different countries available to make this assessment. The first filter we consider is a linear increase of weight between the first and the fifteenth day. The second one is a parabolic growth of the weight and, finally, the third one giving more relevance to only the last three days (hundred times larger than the other twelve days). By comparing the application of the equal weight and the other three filters, we obtain the filter which minimizes the relative error in comparison with the other predictions, see the comparison in Fig. 7(A).

**Figure 7.**
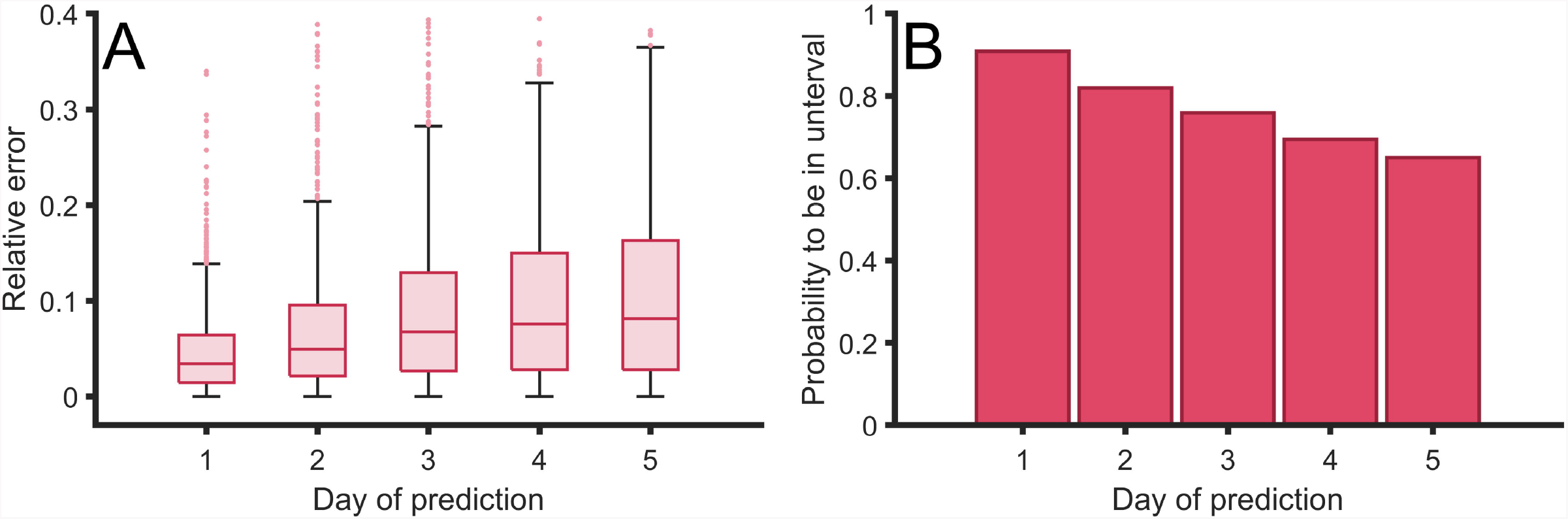
**Error of the predictions with different type of filters.** (A) Relative error between the predictions of the confirmed case for the next five days, in comparison with the actual confirmed cases for four different types of filters: constant values (red), linear increase (orange), parabolic increase (green) and a filteer with large three last values (blue). (B) Probability to obtain the actual real value inside the interval of confidance inside the error bars for the next five days using the same four filters. Light bars to the probability to be found inside confidence interval using each filter confidance intervale, dark bars show probability to be found inside confidence interval using first filter confidance intervale to be able to compare among the different filters. As different filters have different confidence intervals size, although they have the same significance level of α = 0.01.

Although the comparison among the fours procedures, see Fig. 7, shows relative small differences, this statistical study shows a better performance of the last filter, which takes into account larger weights of the last three values of the data. The performance for such filter is particularly better when the epidemic approaches the values of the peak of new cases.

The average of the relative error decreases with the asymmetry of the type of filter we employ, see Fig. 7(A). The filter with higher weights in the last three events presents a better performance in comparison with the three other filters employed. It is important to note that we have also checked other filters with more weight in the last single event or the last two events and the results were less accurate.

We obtain similar results if we evaluate the probability of success of the predictions of each filter, see Fig. 7(B). Light bars in such figure show the success using the error bars obtained from the mean square method adapted to each of the filters. Note that the error bars, or confidence interval, of each method, may be different and therefore is may affect the probability of success basically because it produce larger confidence intervals. To systematically compare the four methods we employ the confidence interval of the original method to the mean values obtained in the other filters. Note that with such definition, the dark and light bars for the first method overlap. We also observe a better performance in the increasing of the asymmetry of the filter and as in the previous comparison the method focused in the last three values maximizes the probability of success.

### Long-term predictions can be obtained from Gompertz model

The use of a phenomenological function facilitates the projection to the future of the trend in comparison with other methods which evaluates in the vicinity of the last day. Although the only relatively reliable predictions in such a complicated problem are short-term predictions, we can however address relevant questions like the final value of total cases of parameter *K*, predictions of the peak or maximum of new cases or the time needed to arrive to the 90% of the total cases. To obtain such long term predictions we employ the whole data set for each country to unveil the trend of the whole dynamics.

We calculate daily the parameter values of the fitting function described above and the evolution of the parameter *K* for different countries together with two characteristic times of the epidemic. See two examples, Spain and Italy, in Fig. 8, for the value of *K*, t_p_ and 90%K in other countries in Europe see, respectively, supplementary Fig. S3 Fig, supplementary Fig.S4 Fig, and supplementary Fig.S5 Fig. The predictions begin with large uncertainty however the values converge to the actual value systematically to the three predictions. The interval of confidence also reduces with time, although there are systematic fonts of errors not addressed by the interval. The main differences between Spain and Italy in the Fig. 8 are the large errors bars of Spain at the beginning of the evolution, because the delay on the epidemic phase of both countries at March 9th, when the graphic begins. While in Italy the epidemic was fully developed, in Spain the epidemic was at the initial phase with an exponential growth.

**Figure 8.**
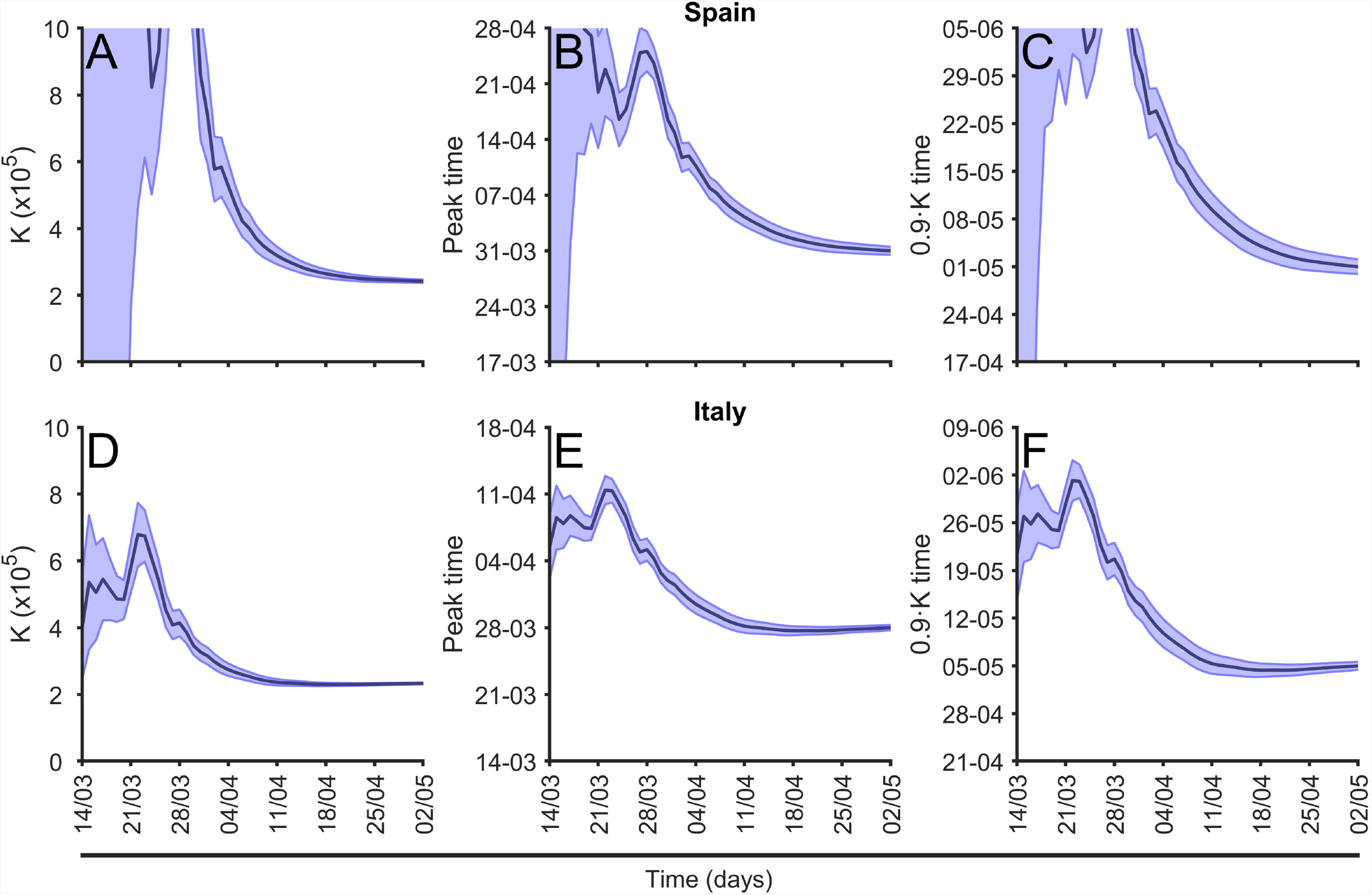
**Evolution of the long-term predictions.** (A) Evolution of the final total number of cases, *K*, prediction; (B) evolution of prediction of the time for the peak of maximum new cases prediction, see eq.(6) and (C) evolution of the arrival to the 90% of total cases time between March 14th and May 2nd in Spain. (D) Evolution of the final total number of cases, *K*, prediction; (E) evolution of prediction of the time for the peak of maximum new cases prediction, see eq.(6) and (F) evolution of the arrival to the 90% of total cases time between March 14th and May 2nd in Italy.

Using the method described above we can compare the three predictions shown in Fig. 8 for all the countries in Europe for a particular date, see this comparison in Fig. 9. For the two temporal comparisons note that actually the dates for the peak for some countries had been already passed in the moment where the evaluation was done. However it is actually not always clear the actual moment when the country is passing the peak. On the other hand, for the comparison among the different countries in Europe with very different demographics, we have used the incidence of the epidemic, evaluated as the number of cases per 10^5^ inhabitants. In this graphic we compare with the actual, at May 2nd 2020, phase of the epidemic in each country [22]. While some of the countries are close to the final number of cases, there are some countries still at the initial phase of the epidemics with very large growth, which predicts large incidence rates. This is the example of United Kingdom.

**Figure 9.**
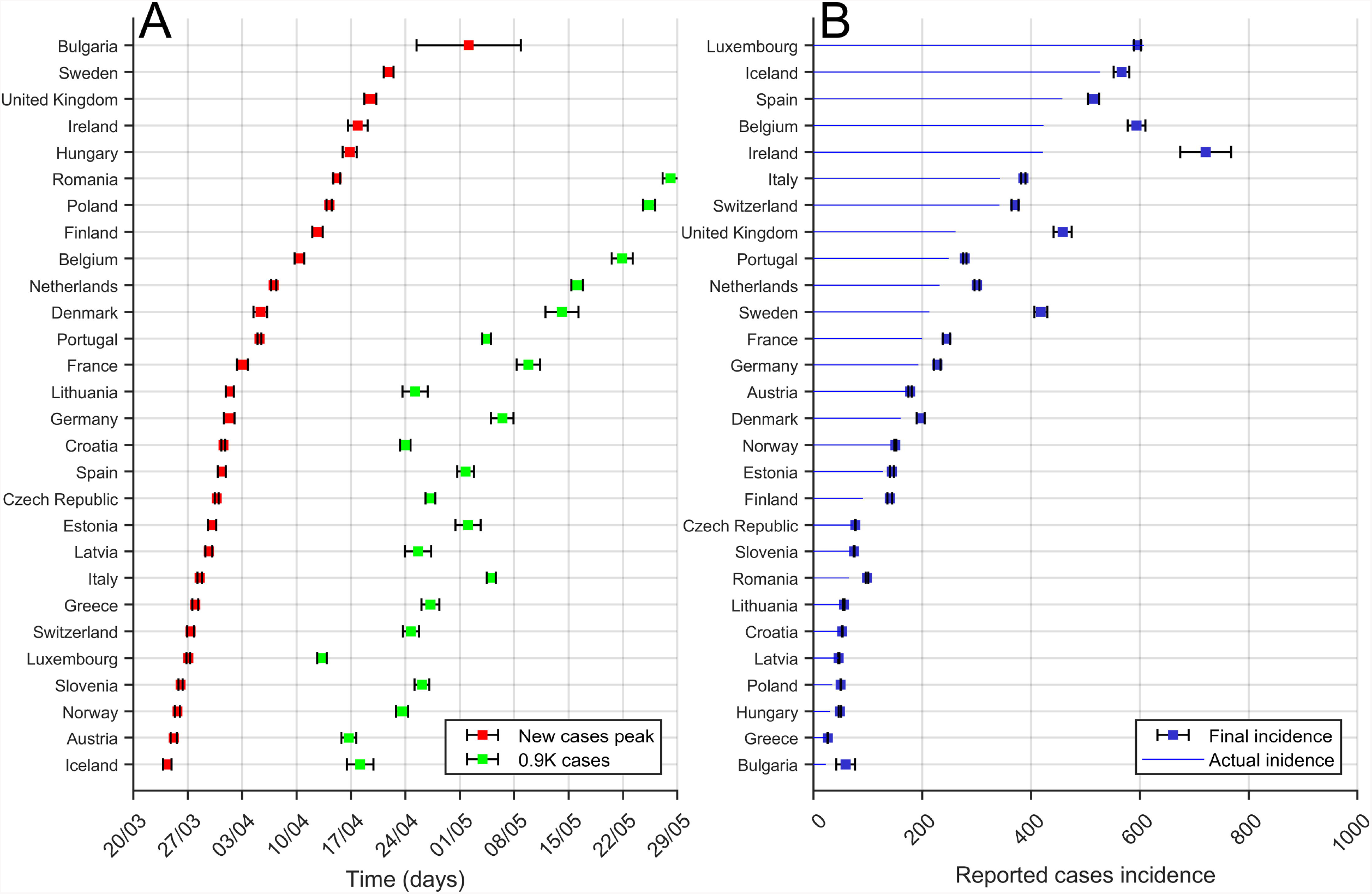
Comparison of long-term predictions among European Countries.

(A) Time for the peak prediction (red) and the time to arrive to the 90% of total cases (in green) predictions obtained from the last evaluation of the Gompertz function (April 12th 2020) to the evolution of the cumulative cases. Countries are sorted from top to bottom using time to peak time prediction. (B) Final incidence (total cases per 10^5^inhabitants) prediction (blue squares) obtained from the last evaluation of the Gompertz function (April 12th 2020) to the evolution of the cumulative cases (blue line), see procedure in Fig. 8. Error bars correspond to the error obtained from the fit and the corresponding error propagation. Countries are sorted from top to bottom actual incidence.

Note that we have to take such predictions reticently, because they are only approximations assuming some simple premises. New epidemic focuses may change completely the dynamics and the values of final incidence for example. Second waves are not considered in the model and we may treat them as an independent epidemic where probably the numbers have to be reseted.

## Discussion

We have fitted the Gompertz function to the cumulative cases in different regions and countries to be able to infer from the fitted parameters of the model relevant quantities for understanding of the epidemics. On the one hand, we have obtained reliable short-time predictions for the new cases during the subsequent days. These predictions are robust and the percentage of success is around 90% for the next day. On the other hand, the fitting allows to provide some long-term quantities, for example, estimations of the total number of cases or the timing of the peak of new cases.

As an empiric function, Gompertz does not depend on previous knowledge of the system. It is specially useful in situations where there is no deep knowledge of the internal structure of the epidemics and when key properties of the epidemics are not known. Precisely, the lack of knowledge regarding the different pathways of contagion or its dependence on social measure makes the fitting of quantitative predictable model impossible. Complex models with a lot of parameters to fit are, in this type of epidemics, exercises trying to unveil possible scenarios, but never a real quantitative tool. No model can predict the reaction of the population to a particular measure, nor even properly assess the parameters of mobility when even basic immunity questions remain unsolved. This is what makes our results about the large degree of confidence in terms of short-term predictions of the evolution of the Covid-19 epidemics so important. Our work has important ramifications since it can predict and, at the same time, assess, changes in the dynamics of the pandemic. The prediction procedure adapts to changes in any of the structural properties of the system. Changes in the diagnose testing needed to detect a case, in social measures or in the way of counting cases just introduce variation in the model that fade away as the new properties emerge again. We have clearly shown in this paper that this changing structure is properly captured with the decreasing nature of the growth given by parameters µ_0_ and a, and the final number of cases K. The highly complex and unknown nature of key elements of the epidemics does not prevent us prediction its evolution in the short-term and to assess the control, or lack of thereof, of the epidemic spread.

We conclude that the methodology here presented can be further employed for the evaluation of the epidemic and the control measurements in the next countries where the spread is on its initial stage. We are planning to further collaborate with health institutions in Africa and America to advise them with the predictions of the model for the evolution of the Covid-19 epidemic in these countries.

In such collaboration, the continuous interplay between predictions and results during spreading will bring us to the rethinking of the assumptions of our model and the further improving of the predictions by the introduction of changes and improvements. Further work can be done to improve the prediction process. The results of the fitting might be better if country-wide data is disaggregated for more homogeneous subnational regions. Data shows that in some countries the appearance of different focus produces the formation of different epidemics which under the conditions of strong restriction of the movement can give rise to independent dynamics inside the country. It is more reliable to work with the region information although the number of cases is lower and the fluctuations stronger. However, the main limitation of the regional approach is the lack of detailed data and/or the difference in the protocols and definitions taken by local authorities.

Finally, we would like to note that the use a generic function is an empiric tool to treat future local and global epidemics, as it has been begun recently with other growth functions like Verhust and Richards models [17]. We plan to continuously update the approach employed here to adapt to any special particularity of any new epidemics. Presently, the same data is applied to guide public policy in hospital administrations giving assessment to regional governments regarding the short-term evolution of health needs.

In order to take adequate and precise control measurements politicians need updated information of the epidemics and a clear representation of the phase of the epidemic among several countries or in a particular country of the different regions. The short-time predictions area valuable information of great interest to politicians.

## Data Availability

All data is available

https://wwwwhoint/emergencies/diseases/novel-coronavirus-2019/situation-reports

## Supporting information

**S1 Fig. Cases in different European countries.** The total cases together with the new daily cases with the corresponding fitings obtained from the Gompertz model are shown for a selection of european countries.

**S2 Fig. Evolution of the fitting of parameter *a*.** The dynamics of the fitting of the parameter a obtained from fiting from the Gompertz model is shown for a selection of european countries.

**S3 Fig. Evolution of the fitting of parameter *K*.** The dynamics of the fitting of the parameter *K* obtained from fiting from the Gompertz model is shown for a selection of european countries.

**S4 Fig. Evolution of the fitting of parameter *t_p_*.** The dynamics of the fitting of the parameter t_p_ obtained from fiting from the Gompertz model is shown for a selection of european countries.

**S5 Fig. Evolution of the fitting of parameter 90%*K*.** The dynamics of the fitting of the parameter 90%*K* obtained from fiting from the Gompertz model is shown for a selection of european countries.

## Acknowledgments

CP, PJC and MC received funding from La Caixa Foundation (ID 100010434), under agreement LCF/PR/GN17/50300003; PJC received funding from Agència de Gestió d’Ajuts Universitaris i de Recerca (AGAUR), Grup Unitat de Tuberculosi Experimental, 2017-SGR-500; CP, DL, SA, MC received funding from Ministerio de Ciencia, Innovación y Universidades and FEDER, with the project PGC2018-095456-B-I00. This work has been also partially funded by the European Comission – DG Communications Networks, Content and Technology through the contract LC-01485746.

## Notes

### Competing Interest Statement

The authors have declared no competing interest.

